# Impact of Restrictions on Parental Presence in Neonatal Intensive Care Units Related to COVID-19

**DOI:** 10.1101/2020.07.22.20158949

**Authors:** Ashley Darcy-Mahoney, Robert D. White, Annalyn Velasquez, Tyson S. Barrett, Reese H. Clark, Kaashif A. Ahmad

## Abstract

**Objectives:** To determine the relationship between the emergence of COVID-19 and neonatal intensive care unit (NICU) family presence as well as how NICU design affects these changes.

**Study Design:** A cross-sectional survey from April 21 to 30, 2020. We queried sites regarding NICU demographics, NICU restrictions on parental presence, and changes in ancillary staff availability.

**Results:** Globally, 277 facilities responded to the survey. NICU policies preserving 24/7 parental presence decreased (83% to 53%, p<0.001) and of preserving full parental participation in rounds fell (71% to 32%, p<0.001). Single family room design NICUs best preserved 24/7 parental presence after the emergence of COVID-19 (single family room 65%, hybrid-design 57%, open bay design 45%, p=.018). In all, 120 (43%) NICUs reported reductions in therapy services, lactation medicine, and/or social work support.

**Conclusions:** Hospital restrictions have significantly limited parental presence for NICU admitted infants, although single family room design may attenuate this effect.

## Background

Approximately 8.4% of United States newborns are admitted into neonatal intensive care units (NICUs) every year [1]. For extremely premature infants or those with major congenital anomalies, these admissions can last several months. Extended intimate contact of a newborn with its parents, both mother and father, has been shown to have physiological and psychological benefits to both newborn and parents [2]. This contact, often referred to as skin-to-skin or kangaroo care, provides many familiar sensory inputs such as the mother’s voice, taste, and smell that cannot be replicated by others and improves physiological stability to the preterm infant [3]. This contact establishes a lasting bond that promotes both neurodevelopment in the infant [4] and enhanced mental health in the mother [3]. In addition to causing significant anxiety, acute stress, and post-traumatic stress for parents, admission to the NICU signifies a disturbance in the maternal-infant bond, which can be detrimental to the development of the newborn [4]. The introduction of family centered care and allowing 24-hour parental presence, especially in the setting of single-family rooms, may help increase parental presence, ameliorate some of these effects [5] and improve developmental outcomes [6].

In response to the emergence of Corona Virus Disease 2019 (COVID-19), the Centers for Disease Control and Prevention (CDC) issued a sequential series of recommendations. These included closure of elementary, middle and high schools on 3/12/2020 [7], screening of visitors entering health care facilities on 3/20/2020 [8], and implementation of social distancing on 4/4/2020 which included working from home when possible [9]. While severe illness appears to be most prevalent in adults, transmission by asymptomatic adults and children may be widespread [10, 11]. Multiple media reports have documented prohibition of visitation for hospitalized adults, including when in extremis, in attempts to limit disease transmission [12, 13].

The tension between protection of infants and caregivers from serious infection and the desire to maximize the developmental outcome of newborns forces consideration of several options for restricted interaction, none of them optimal. While neonatal COVID-19 disease appears relatively uncommon, the risk-benefit calculation for restricting NICU access remains unclear. Further, no published data exist regarding the timing of implementing hospital and NICU entry restrictions or the extent of those restrictions after the emergence of COVID-19. We surveyed hospital sites globally to better understand the timeline and rigidity of hospital and NICU entry restrictions. Additionally we queried sites regarding new limitations in ancillary personnel such as therapy services and lactation consultants. We hypothesized that the availability of a large number of private (“single-family”) rooms in a NICU would decrease the likelihood that the most restrictive parental presence policies would be considered necessary, when compared to NICUs in which most or all of the beds were in a multi-bed rooms.

## Methods

We performed a cross-sectional survey of global NICUs, with a focus on the United States, to determine hospital and NICU entry policies prior to and during the COVID-19 pandemic. The 21-item survey was built of both closed and open response items requesting information on policies both before and during the spread of COVID-19 (Supplemental Figure 1). In particular, we requested information regarding location, type of hospital, NICU design, hospital and NICU entry policies, parental presence, and reductions in support staff. The survey was distributed through multiple venues from April 21 through April 30, 2020. These included an e-mail to the American Academy of Pediatrics (AAP) Section of Neonatal-Perinatal Medicine listserv of NICU medical and quality improvement directors, MEDNAX practice medical directors, and to the MEDNAX Neonatology Forum.. In parallel, we released the survey on Twitter, LinkedIn, Facebook, and group chats of neonatologists and neonatal nurse practitioners. In all venues we requested a response from the NICU medical or nursing director.

We included only those responses with at least completion of the first two sections of the survey (NICU demographics and visitation practices) and excluded multiple responses from the same NICU. To evaluate duplicate responses, we examined NICU zip code and baseline demographics. We determined *a priori* to include the first fully completed survey from any NICU or, in the case of multiple partially completed surveys, the first submitted response. While the stated preference was for NICU medical or nursing directors to respond to the survey, due to the methods of distribution this could not be controlled for.

The primary outcomes of interest were the presence of policies allowing 24-hour parental presence in the NICU and allowing full parental participation in NICU rounds. Secondary outcomes of interest included screening policies related to hospital entry and the impact of COVID-19 on NICU staffing availability. The study was approved by the Methodist Healthcare Institutional Review Board, San Antonio, TX.

### Data Analysis

Two main approaches were used to analyze the data: sample descriptive statistics and comparisons of policies from before the spread of COVID-19 to after the start of the spread. For descriptive statistics, we used counts and percentages, presented in tables and figures. For policy comparisons, given the survey responses were all categorical, we used a series of McNemar’s tests for all pre-post comparisons and Chi-square tests for comparisons between NICU designs.

To adjust for the multiple hypothesis tests, we used a false discovery rate adjustment throughout this manuscript [14]. Analyses were performed in SPSS version 26 and R version 3.6.2.

## Results

A total of 339 responses were received. Of these, 56 were excluded for incompletion of at least the first two survey pages and 6 excluded for duplicate NICU responses, leaving 277 surveys for analysis. Respondents most frequently learned of the survey through AAP and MEDNAX email requests (83%) and responding NICUs were largely in the United States (91%) with Texas, California, and Florida NICUs comprising 33% of the total cohort (Table 1, Supplemental Figure 2a). The characteristics of the responding NICUs are described in Table 1.

**Table 1.**
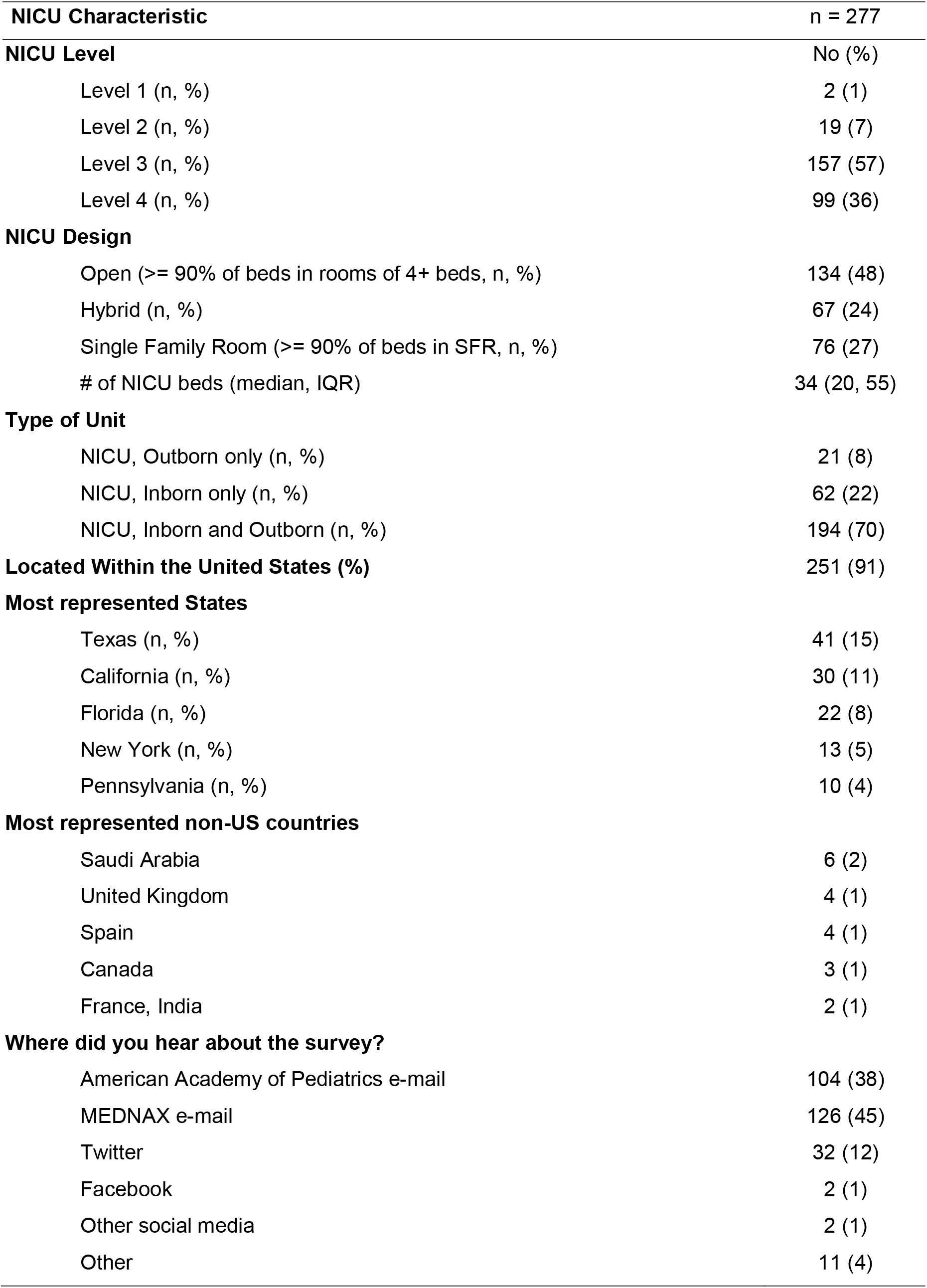
Demographics of Responding NICUs.

| **NICU Characteristic** | **n = 277** |
| --- | --- |
| **NICU Level** | **No (%)** |
| Level 1 (n, %) | 2 (1) |
| Level 2 (n, %) | 19 (7) |
| Level 3 (n, %) | 157 (57) |
| Level 4 (n, %) | 99 (36) |
| **NICU Design** |  |
| Open (>= 90% of beds in rooms of 4+ beds, n, %) | 134 (48) |
| Hybrid (n, %) | 67 (24) |
| Single Family Room (>= 90% of beds in SFR, n, %) | 76 (27) |
| # of NICU beds (median, IQR) | 34 (20, 55) |
| **Type of Unit** |  |
| NICU, Outborn only (n, %) | 21 (8) |
| NICU, Inborn only (n, %) | 62 (22) |
| NICU, Inborn and Outborn (n, %) | 194 (70) |
| **Located Within the United States (%)** | **251 (91)** |
| **Most represented States** |  |
| Texas (n, %) | 41 (15) |
| California (n, %) | 30 (11) |
| Florida (n, %) | 22 (8) |
| New York (n, %) | 13 (5) |
| Pennsylvania (n, %) | 10 (4) |
| **Most represented non-US countries** |  |
| Saudi Arabia | 6 (2) |
| United Kingdom | 4 (1) |
| Spain | 4 (1) |
| Canada | 3 (1) |
| France, India | 2 (1) |
| **Where did you hear about the survey?** |  |
| American Academy of Pediatrics e-mail | 104 (38) |
| MEDNAX e-mail | 126 (45) |
| Twitter | 32 (12) |
| Facebook | 2 (1) |
| Other social media | 2 (1) |
| Other | 11 (4) |

Across the United States, the timeline for implementing state-wide stay at home orders did not begin until the second half of March [15]. However, hospital and NICU entry policies began to change in the first week of January with a rapid increase in hospitals adopting policy changes throughout March, the majority prior to the issuance of CDC guidance (Supplemental Figure 3). At the state level, the mean date for changing hospital entry policies varied significantly. Hospitals in 4 states averaged implementation of hospital restrictions as early as February and 3 states as late as April (Supplemental Figure 2b). The first international NICU restricted entry in January, followed by 21 units in March, and 5 in April. Overall, 184 (66%) NICUs reported that their new policies during the COVID-19 pandemic were broadly more restrictive than the customary policies implemented during the winter influenza / respiratory syncytial virus season.

Changes in overall hospital entry screening policies became widespread during the COVID-19 pandemic (Table 2). These included significant increases in physical temperature checks as well as screening questions regarding travel history, fever, and illness for hospital entry.

**Table 2.**
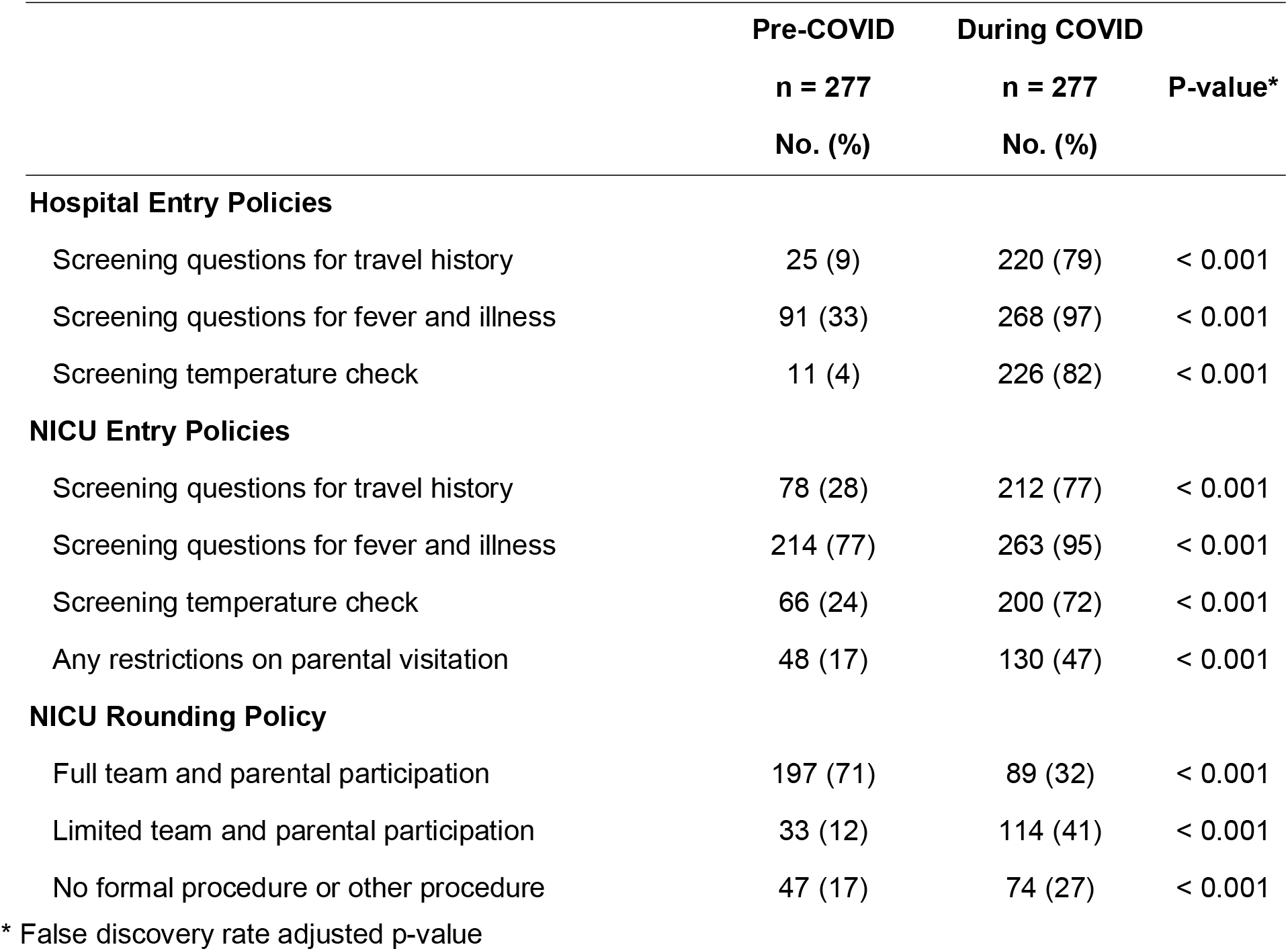
Hospital and NICU Policy Changes During the COVID-19 Pandemic.

Hospitals implemented parallel changes affecting NICU entry after the emergence of COVID-19 (Table 2). NICUs broadly implemented screening questions for travel history (76.5%), fever and illness (94.9%) as well as physical temperature checks (72.2%). The number of NICUs allowing 24-hour parental presence in the NICU decreased significantly (83% to 53%, p < 0.001). While full team and parental participation in NICU rounds predominated prior to the COVID-19 pandemic, this became significantly less common during the pandemic (71.1% vs.32%, p < 0.001, Table 2). The number of states with over 80% of responding NICUs allowing full team and parental participation in NICU rounds fell from 20 (50%, Supplemental Figure 2c) to 3 (8%, Supplemental Figure 2d).

Within the 130 NICUs that restricted parental presence during the COVID-19 pandemic, we found a variety of policy measures described (Supplemental Figure 4). This included 7 NICUs (5%) excluding all parental presence, 2 (2%) of which would not allow parents to enter even with their infant in extremis. Most NICUs restricting parental presence only allowed one parent at the bedside at any time (85%) and a minority (25%) required families to choose a single parent to be allowed into the NICU for the entire hospital stay.

We examined the influence of NICU design (all open bay, single family room, or hybrid) on NICU entry and NICU rounding policies. All NICU design types implemented significant increases in routine screening measures after the COVID-19 outbreak and had significant decreases in 24-hour parental presence, but these restrictions were less in single family room NICUs (84% to 64%, absolute difference 20%, p = .009) compared to hybrid (91% to 57%, absolute difference 34% p < 0.001) and open bay units (78% to 45%, absolute difference 33%, p < 0.001, Table 3). While 24-hour unrestricted parental presence was not significantly different between all NICU designs prior to the COVID-19 pandemic, we found significant differences during the pandemic (single family room 65%, hybrid-design 57%, open bay design 45%, p =.018).

**Table 3:**
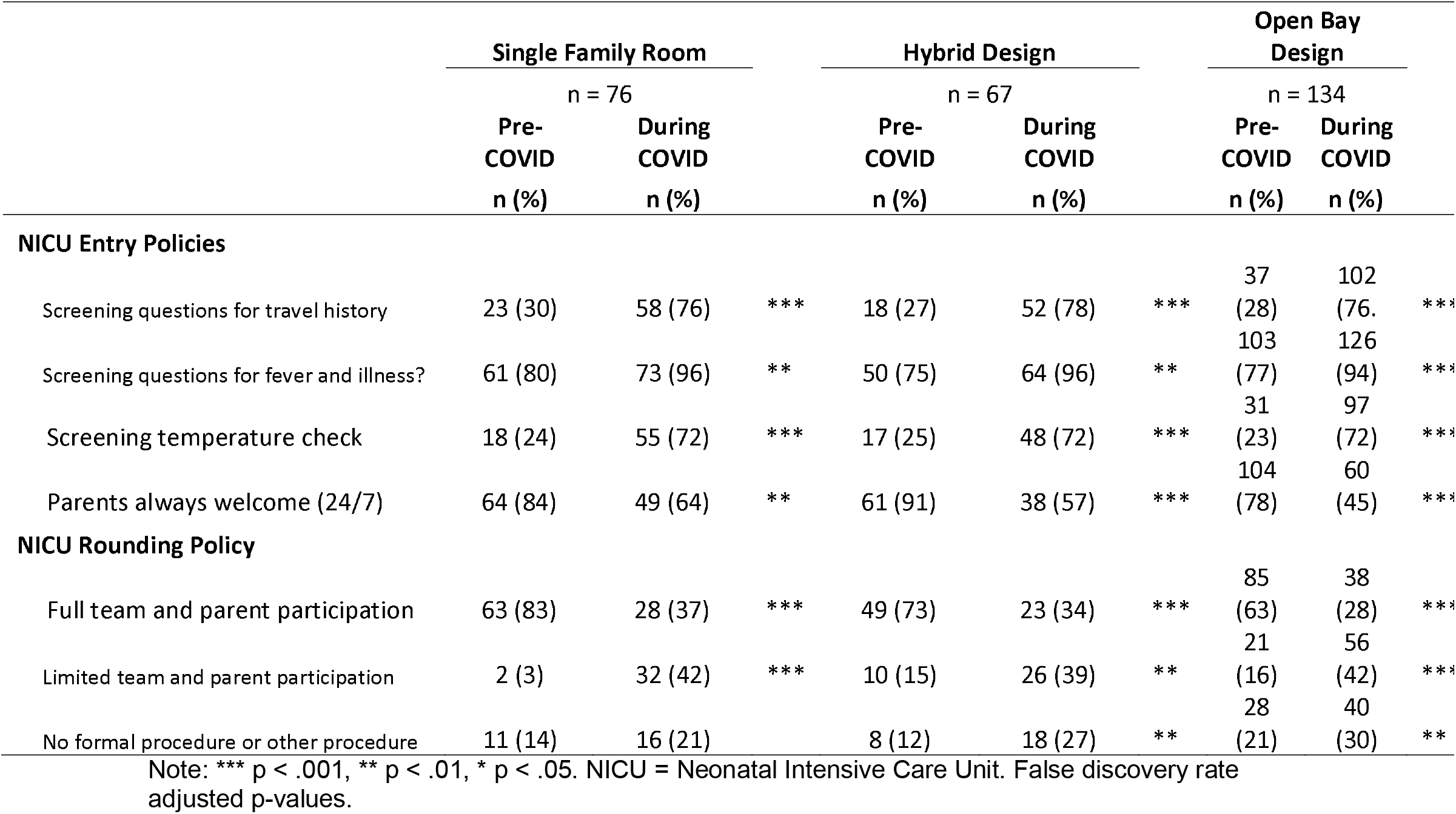
The Effect of NICU Design on the Changes in NICU Entry and Rounding Policies

|  | Single Family Room |  |  | Hybrid Design |  |  | Open Bay Design |  |  |
| --- | --- | --- | --- | --- | --- | --- | --- | --- | --- |
|  | n = 76 |  |  | n = 67 |  |  | n = 134 |  |  |
|  | Pre-COVID | During COVID |  | Pre-COVID | During COVID |  | Pre-COVID | During COVID |  |
|  | n (%) | n (%) |  | n (%) | n (%) |  | n (%) | n (%) |  |
| **NICU Entry Policies** |  |  |  |  |  |  |  |  |  |
| Screening questions for travel history | 23 (30) | 58 (76) | *** | 18 (27) | 52 (78) | *** | 37 (28) | 102 (76) | *** |
| Screening questions for fever and illness? | 61 (80) | 73 (96) | ** | 50 (75) | 64 (96) | ** | 103 (77) | 126 (94) | *** |
| Screening temperature check | 18 (24) | 55 (72) | *** | 17 (25) | 48 (72) | *** | 31 (23) | 97 (72) | *** |
| Parents always welcome (24/7) | 64 (84) | 49 (64) | ** | 61 (91) | 38 (57) | *** | 104 (78) | 60 (45) | *** |
| **NICU Rounding Policy** |  |  |  |  |  |  |  |  |  |
| Full team and parent participation | 63 (83) | 28 (37) | *** | 49 (73) | 23 (34) | *** | 85 (63) | 38 (28) | *** |
| Limited team and parent participation | 2 (3) | 32 (42) | *** | 10 (15) | 26 (39) | ** | 21 (16) | 56 (42) | *** |
| No formal procedure or other procedure | 11 (14) | 16 (21) |  | 8 (12) | 18 (27) | ** | 28 (21) | 40 (30) | ** |

Note: \*\*\* p < .001, \*\* p < .01, \* p < .05. NICU = Neonatal Intensive Care Unit. False discovery rate adjusted p-values.

With respect to parental presence during rounds, prior to the emergence of COVID-19, we found significant differences in NICUs allowing full parental presence in rounds based on NICU design (SFR 83%, hybrid NICUs 73%, open bay NICUs 63%, p = 0.013). However, all NICU design types imposed a significant restriction in parental participation in NICU rounds during the pandemic (Table 3). After the emergence of COVID-19 we found no significant differences in full parental participation in NICU rounds across design types (SFR 37%, hybrid 34%, open bay 28%, p = 0.6).

NICUs reported significant changes in staffing or outcome measures temporally associated with the COVID-19 pandemic. These included 28% of sites reducing nursing staff hours, 28% reducing physical (PT), occupational (OT), and/or speech therapy (ST) and 23% delaying non-urgent procedures (most commonly gastrostomy tubes). We also found 21% of NICUs reported decreased lactation consultant visits, 16% restricted donor milk utilization, and 8% reported changes in breastfeeding rates. In all, 120 (43%) NICUs reported reductions in therapy services, lactation medicine, and/or social work support. Social services and lactation medicine offered supplemental telehealth support in 109 (41%) NICUs.

## Discussion

Prohibitions on family presence for critically ill or dying adults as a result of COVID-19 pandemic have been widely reported by the media. Little is known however regarding how COVID-19 has impacted family presence for minors, including the most vulnerable in our population – those admitted to NICUs. These data represent the first report documenting the widespread, rapid, and profound restrictions in hospital and NICU visitation practices secondary to the COVID-19 pandemic. Policy changes began in early January and hospitals rapidly adopted entry limitations throughout March 2020. The result has been a significant shift in family presence for sick infants and of the way parents experience the first phases of their newborn’s lives in the NICU environment. An additional secondary effect of COVID-19 has been delays in elective procedures and the reduction of staff in many NICUs including therapy services, lactation support, and social services -- all important for optimal outcomes. Ultimately, as health care systems attempt to prevent the spread of coronavirus, new policies have led to families interacting very differently with their infants in the NICU — or not at all. These rapidly instituted changes may carry with them the risk of secondary unintended consequences [16].

In this cross-sectional survey of 277 NICUs we found widespread implementation of hospital and NICU screening measures, including decreases in NICUs allowing 24-hour parental presence (83% to 53%). Single family room NICU design may attenuate some restrictions. Private rooms make certain infection control measures easier by providing physical distancing, physical barriers, and separate air supplies [17, 18]. At baseline we found no differences in 24-hour parental presence among various NICU designs. However, we found more NICUs with single-family room design able to maintain 24-hour parental presence and interaction with babies and caregivers than open bay units (64% vs 45%). This clinically meaningful preservation of parental presence occurred at a time when many other limitations were imposed, including limiting parental participation in NICU rounds.

We find that restrictions in NICU parental presence have been widely adopted and that the ability for parents to participate in aspects of shared decision making such as family centered rounds have diminished significantly. Family-centered rounds (FCRs) are multidisciplinary rounds that occur at the patient’s bedside, in the presence of patients and family members, and integrate patient and parent perspectives and preferences into clinical decision-making. Prior to the COVID-19 pandemic, 71% of NICUs allowed full team and parental participation in NICU rounds, falling to 32% during the pandemic. Family-centered care has been recommended by both the Institute of Medicine [19] and AAP [20] as key to improving the quality and safety of health care. Recommendations include conducting attending rounds in patients’ rooms with family members present and ensuring that decisions on care plans for patients should be made only after such rounds, to incorporate family involvement in decision making. Identified benefits of FCR include improved patient satisfaction, communication, discharge planning, medical education, and patient safety [21]. It has also been suggested that family involvement in care can decrease family stress while improving patient’s outcomes [22, 23]. The abrupt restrictions in parental presence and family participation in rounds necessarily also disrupts the ability to provide family centered care. The impact of these changes on parental stress, patient safety, and patient outcomes require further investigation.

Prior infectious outbreaks, such as Severe Acute Respiratory Syndrome (SARS), resulted in increased mental health disorders, including post-traumatic stress disorder, anxiety, and depression [24]. Data from Wuhan, China demonstrates a similar increase in psychopathology, with some reports suggesting that women may have an increased burden of mental health disorders in the current pandemic [25, 26]. Pregnancy has known associations with significant emotional distress under normal circumstances, and women’s mental health in the perinatal period is significantly affected by access to social support and resources [27, 28]. This raises the concern that pregnant and postpartum women may be particularly vulnerable to the psychosocial hardships caused by the pandemic. Studies have also suggested that higher maternal stress may lead to reductions in warm, contingent caregiving (i.e., caregiver responsiveness), which in turn can affect infant neurodevelopment [29, 30]. The described restrictions on parental presence for ill newborns may further magnify the existing stressors associated with the perinatal period and NICU admission.

An important component of the care for ill newborns, especially those born prematurely, is the developmental team, whose members include disciplines such as OT, PT and ST with involvement from social work, and lactation consultants [31, 32]. Overall, 43% of responding NICUs reported decreases in at least one area of these support services. While some sites offered telehealth options for these services, the efficacy of telehealth in this setting is unknown. In addition to direct patient care, the developmental team plays an important role in educating parents on key techniques to implement on a daily basis both in the hospital and post-discharge [33, 34]. While possibly implemented in an effort to minimize the spread of COVID-19 to vulnerable patients, diminished social work and developmental care services to premature infants may have long-term adverse consequences.

Strengths of this study include responses from a large number of hospitals and NICUs across the United States and around the world. These data are sufficient to provide a granular understanding of the timeline by which hospitals and facilities implemented restrictions and how access became significantly limited even in areas of the hospital with minimal disease burden from COVID-19 such as the NICU. Further, many facilities provided data regarding limitations in support services, providing indications of the secondary effects the COVID-19 pandemic has quickly caused. Limitations include those inherent to survey studies including inability to verify responses, incomplete responses, and inability to ask follow-up questions of respondents. In addition, despite our stated intention to receive responses from NICU medical or nursing directors, the identity of the respondent could not be controlled for. We cannot rule out further evolution of hospital policies after the completion of the survey window. Lastly, due to the distribution platforms used for this survey, a denominator and thus response rate could not be determined.

## Discussion

In summary, we report dramatic and rapid changes in hospital and NICU entry policies related to COVID-19. These changes have significantly affected parental presence for NICU admitted neonates, although single family room design may partially attenuate this effect. In addition to these restrictions, limitations in support staff as well as delays in elective procedures for NICU admitted infants are common. In total, the rapid implementation of these sweeping changes may have substantial impact on parental and family well-being and may lead to detrimental effects on neonatal health outcomes. Further investigation regarding the short- and long-term impact of these policy changes is urgently needed.

## Data Availability

After manuscript publication data ca be made available upon request.

## Abbreviations

COVID-19: Corona Virus Disease 2019
NICUs: Neonatal intensive care units
CDC: Centers for Disease Control and Prevention
AAP: American Academy of Pediatrics
PT: Physical Therapy
OT: Occupational Therapy
ST: Speech Therapy
FCR: Family centered rounds
SARS: Severe Acute Respiratory Syndrome
SFR: Single Family Room

## Acknowledgements

We thank Dr. Veeral Tolia for his critical review of this manuscript

## Supplementary Index

**Supplemental Figure 1:**
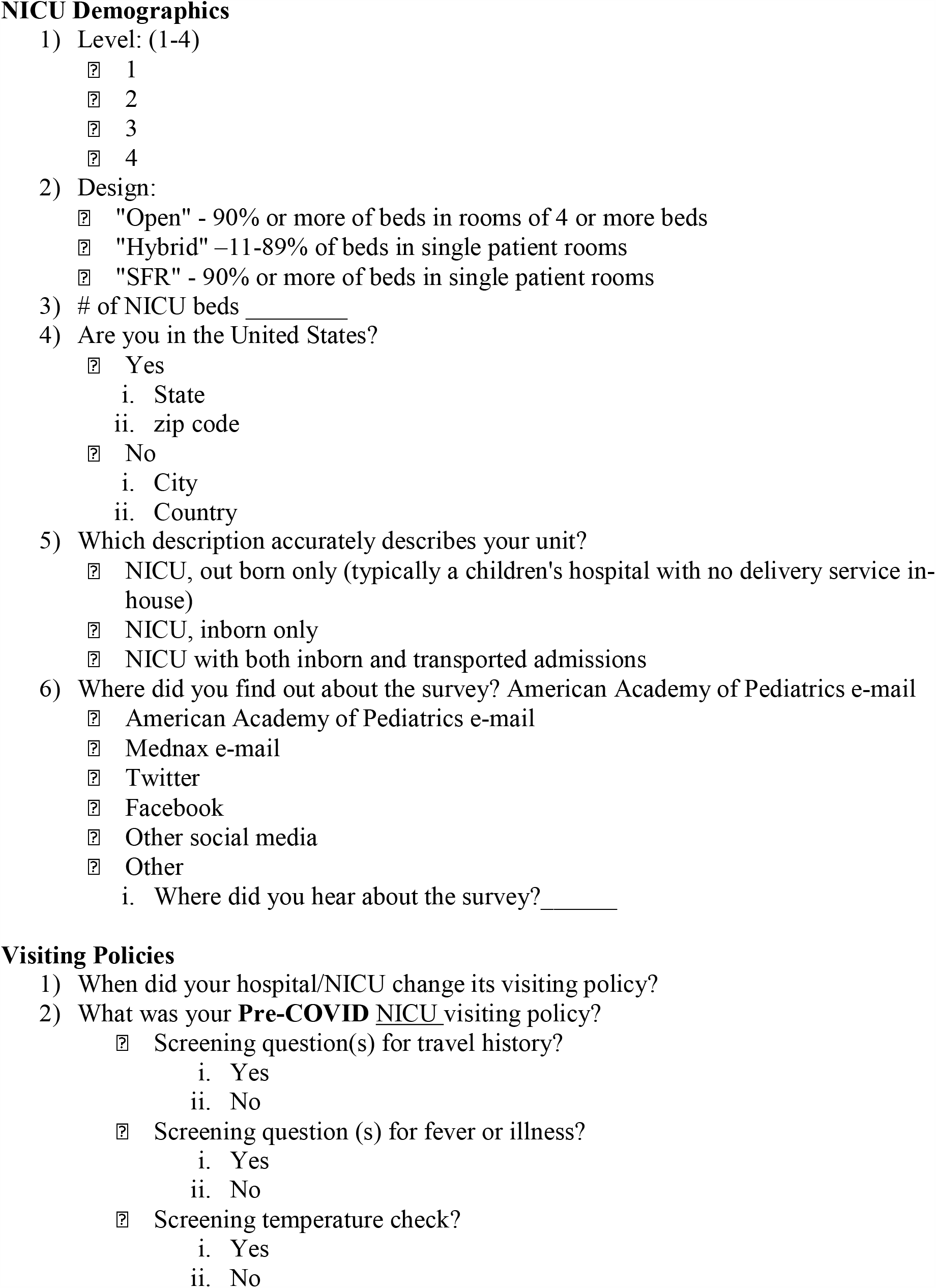
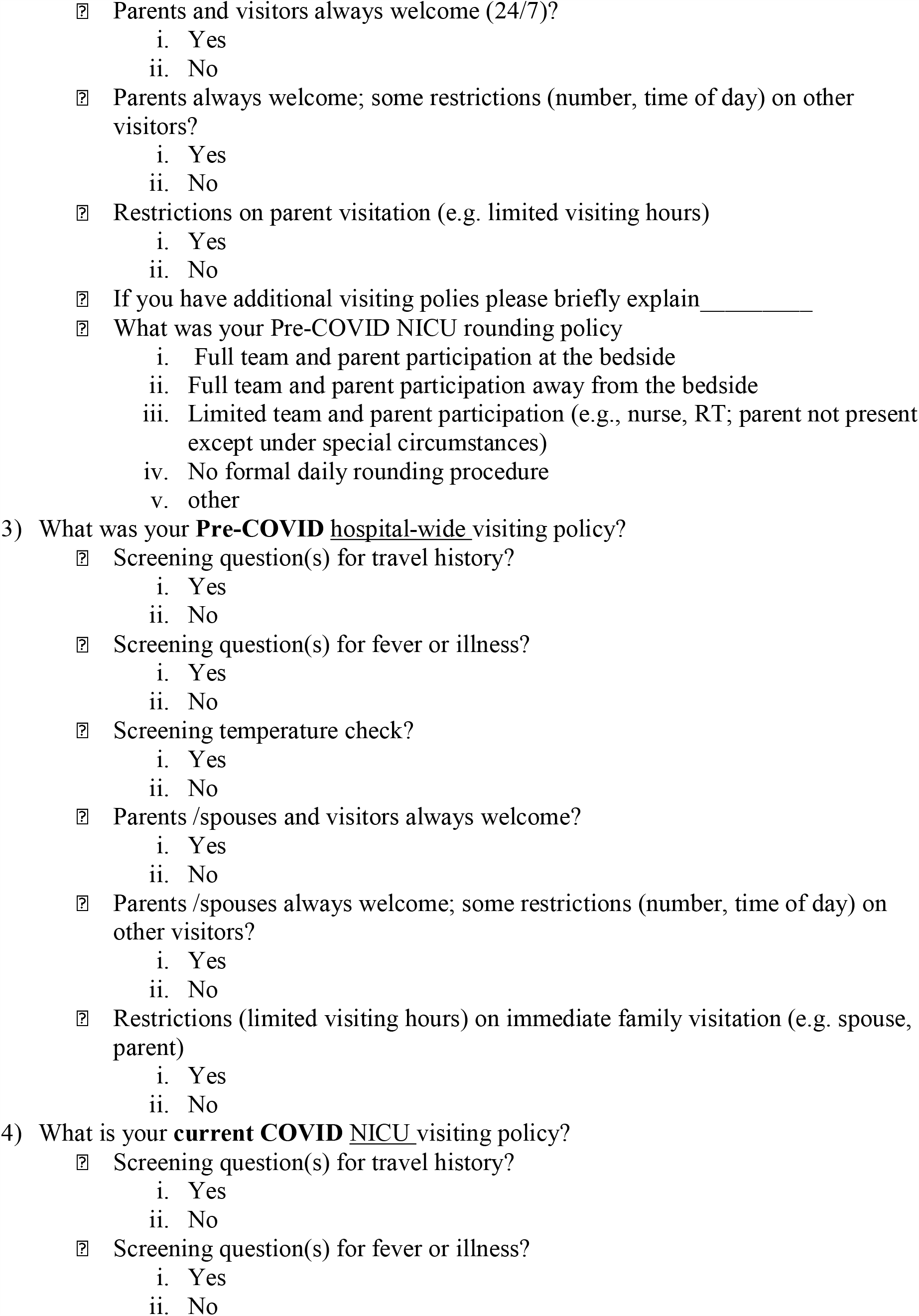
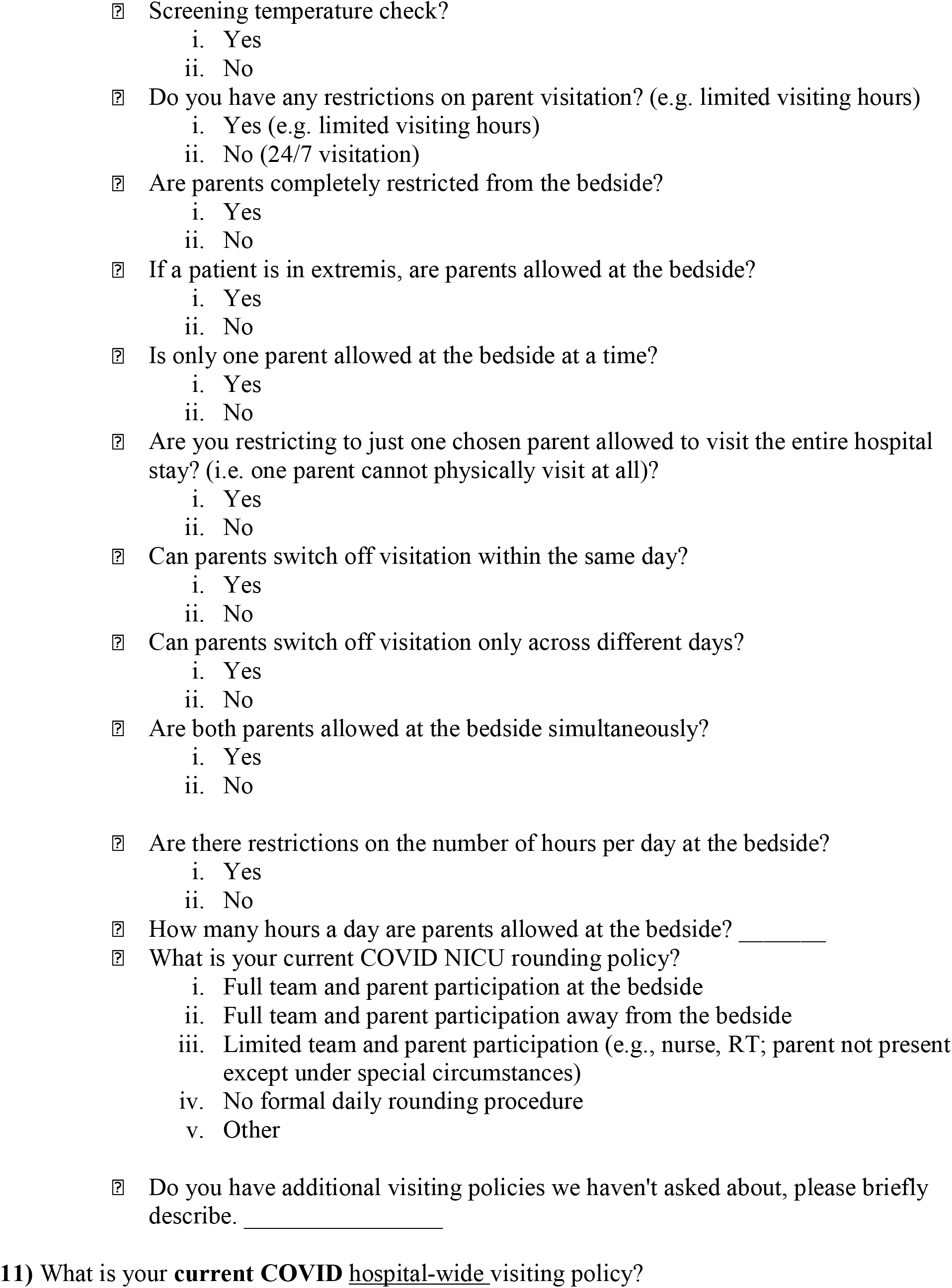
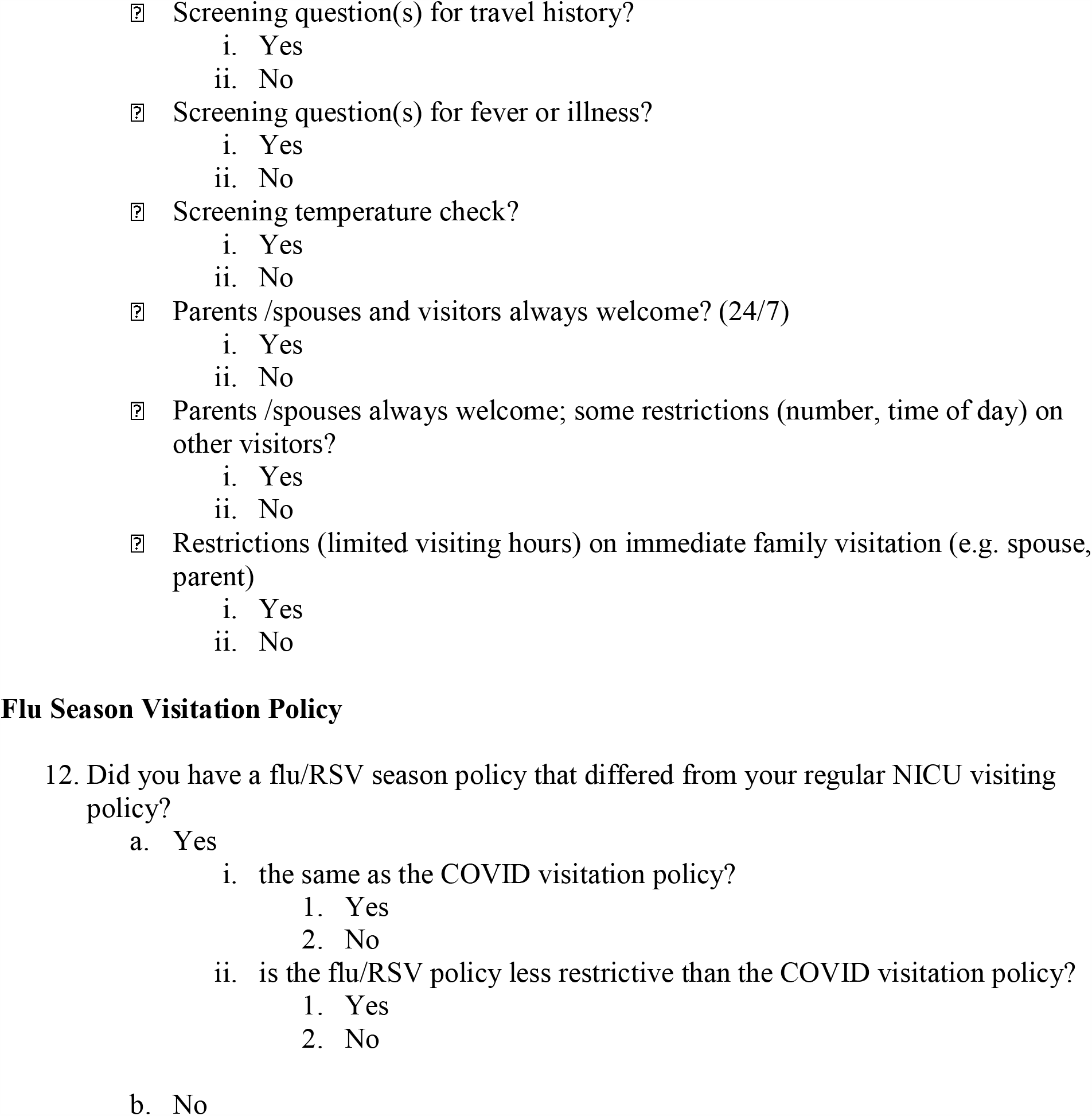
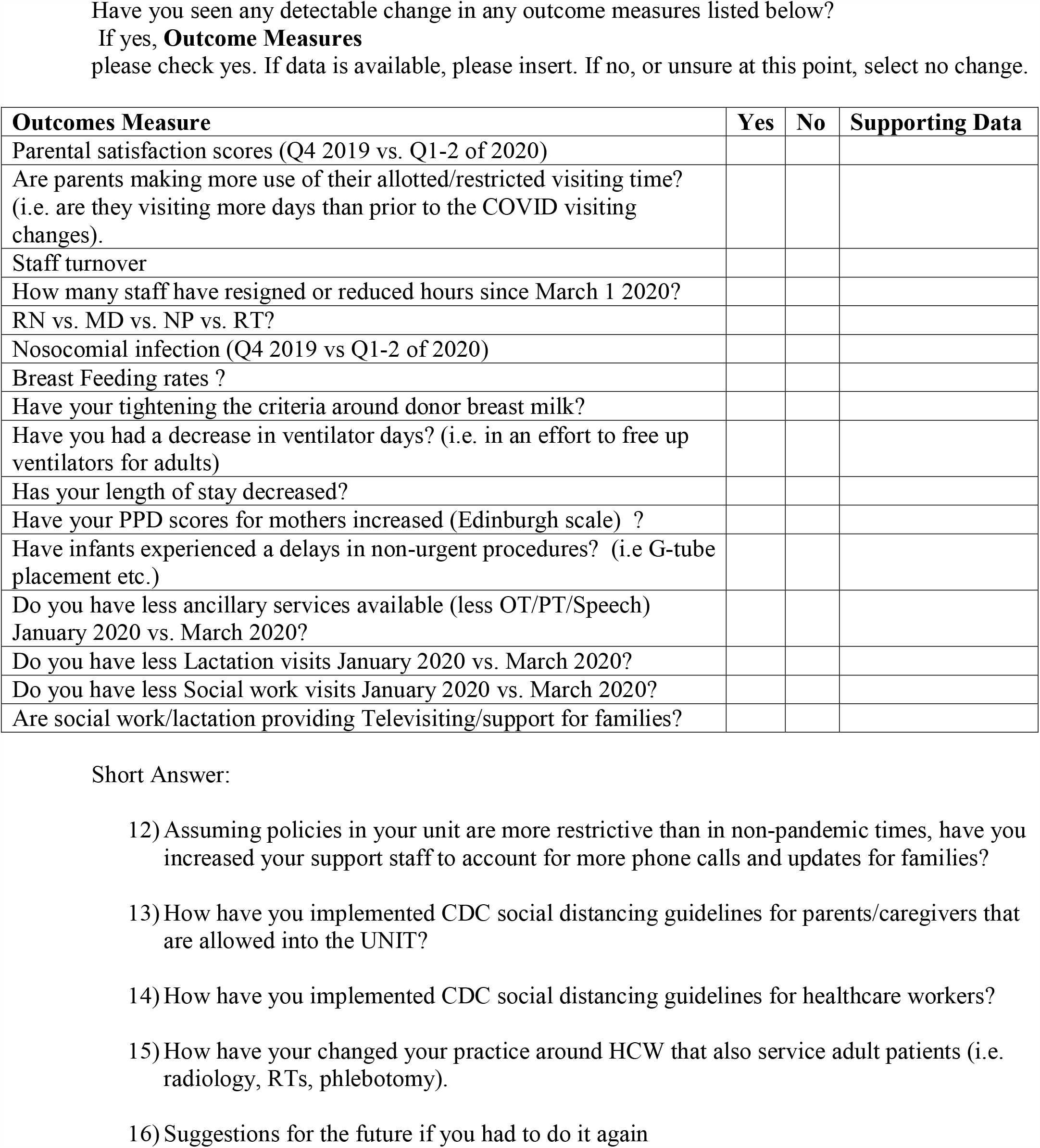
COVID Visitation Questionnaire. The questionnaire was entered into REDCap with branching logic incorporated as appropriate

**Supplemental Figure 2:**
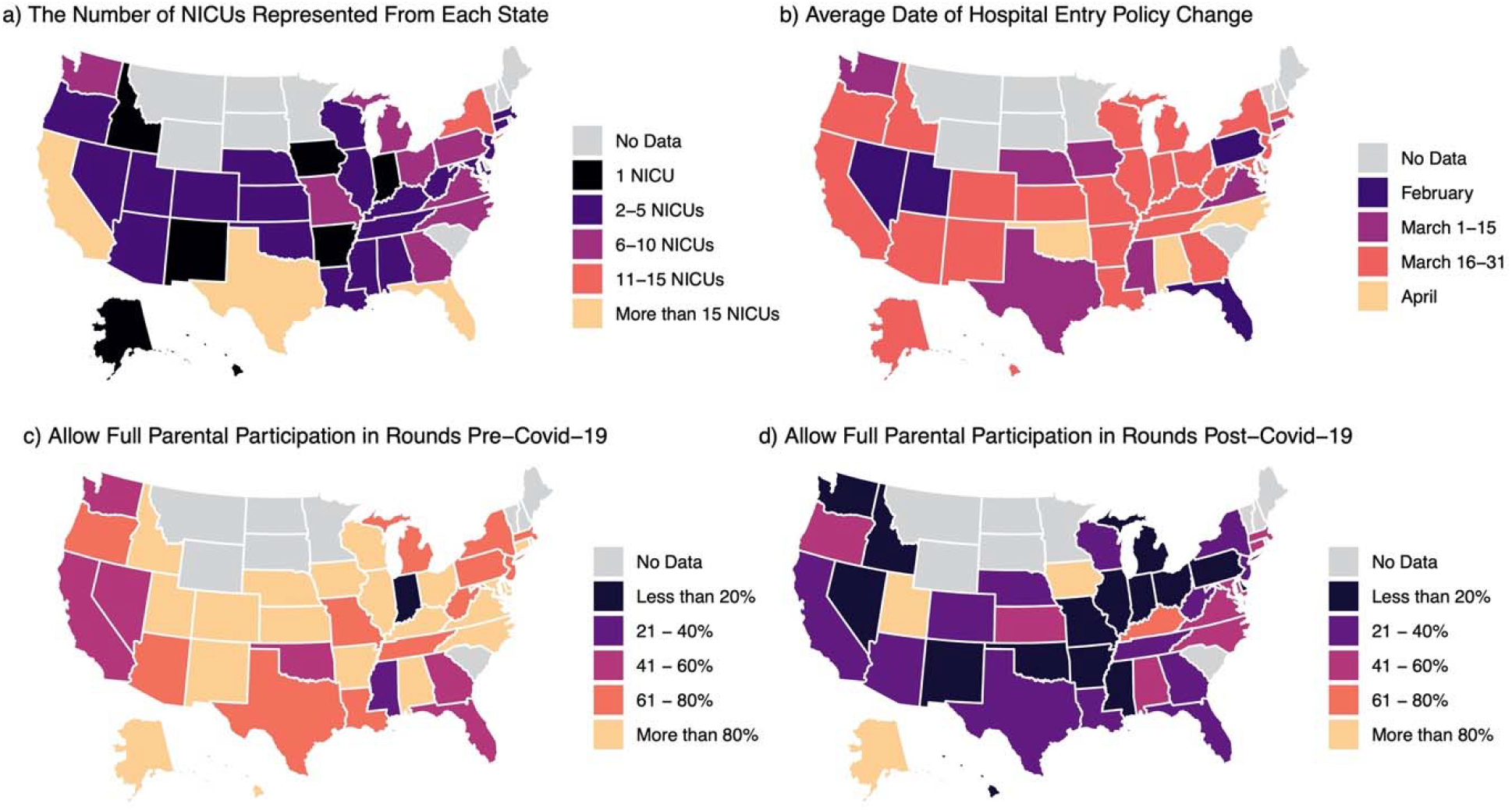
Geographical Distribution of Responding NICUs, Entry Policy Changes, and Level of Parental Participation in Rounds. Responses were received from NICUs in 40 out of 50 US States, with 3 states having more than 15 NICUs respond to the survey (Panel A). The average date of changing hospital entry policies varied by state, with the majority averaging policy change between March 16 and 31 (Panel B). Prior to the onset of the COVID-19 pandemic, permitting full parental participation in NICU rounds was commonplace (Panel C). However, during the pandemic very few states had a majority of NICUs allowing full parental participation in rounds.

**Supplemental Figure 3:**
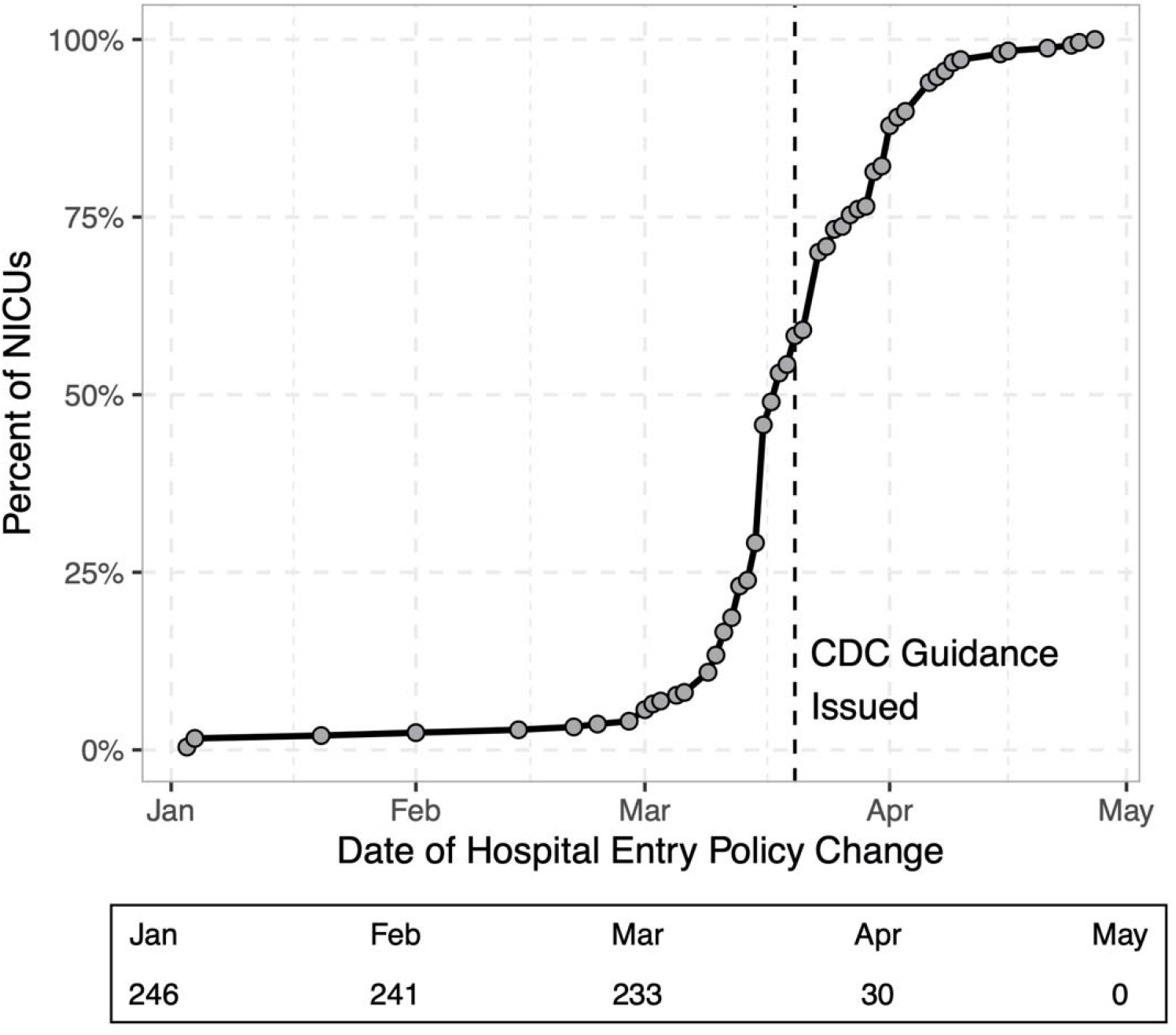
The Cumulative Count of Changes in Hospital Entry Policies by Calendar Date. Very few hospitals altered their entry dates in January or February, 2020. However this rapidly changed in March with most NICUs changing their policy prior to the issuance of specific guidance from the Centers for Disease Control and Prevention (CDC) on March 20, 2020.

**Supplemental Figure 4:**
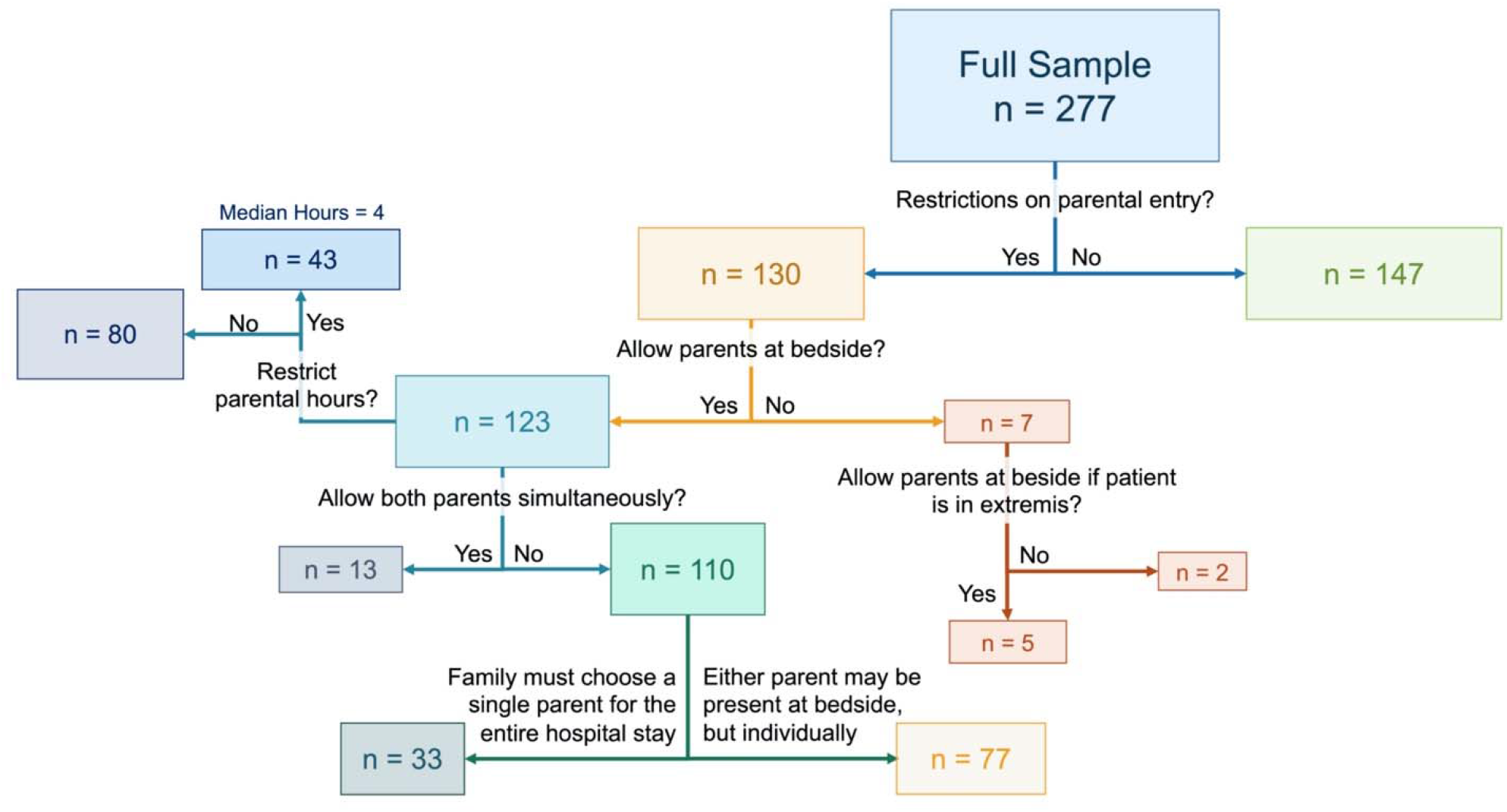
Flow Diagram of Restrictions to Parental Presence in the NICU After the Emergence of Covid-19. Overall, 130 of 277 NICUs (47%) had restrictions to NICU parental in the NICU during the Covid-19 pandemic. Of these, most allowed only a single parent at the bedside and 33 NICUs required families to choose a single parent for the entire hospital stay. Prohibition of parental presence in the NICU was rare but occurred at 7 sites (3%).

## Notes

### Competing Interest Statement

The authors have declared no competing interest.

### Funding Statement

No external funding was received for this work.

### Author Declarations

Methodist Healthcare IRB

